# Adherence to a modified DASH diet, intake of key food groups, and infertility in women and men: the Norwegian Mother, Father and Child Cohort Study

**DOI:** 10.64898/2026.10.21.26366136

**Authors:** Álvaro Hernáez, Ida Henriette Caspersen, Thea Grindstad, Mari Mohn Paulsen, Sara Castro-Barquero, Marta H. Hernández, Maria Christine Magnus

**Author notes:** **Corresponding author:** Álvaro Hernáez, PharmD, PhD, Hospital del Mar Research Institute (HMRIB), Carrer Doctor Aiguader 88, 08003, Barcelona (Spain).

## Abstract

**Background & Aims:** Diet is a modifiable exposure that may contribute to reproductive health. While most previous studies focused on selected nutrients in relation to fertility treatment outcomes or semen parameters, large cohort data addressing overall diet quality and clinical infertility in both women and men remain limited. We aimed to assess how adherence to a modified Dietary Approaches to Stop Hypertension (DASH) dietary pattern and intake of key food groups relate to infertility in women and men.

**Methods:** We included 59,218 women and 26,866 men participating in the Norwegian Mother, Father and Child Cohort Study (MoBa). Diet was assessed using dietary questionnaires and summarized using a modified DASH score comprising fruits, vegetables, nuts/legumes, whole grains, low-fat dairy, sweet beverages, red and processed meat, and a snack/fast food proxy. Fish was also assessed. Infertility was defined as having tried to conceive for at least 12 months or using assisted reproductive technologies. Logistic regression models were fitted separately in women and men, adjusting for age, education, body mass index, smoking, and number of offspring. Food-group analyses were evaluated using a multiple-comparison threshold of *p*-value <0.0167, derived from Horn’s parallel analysis.

**Results:** A higher modified DASH score was associated with lower odds of infertility in women (adjusted OR per 1-SD increment: 0.97, 95% CI 0.95 to 0.995) and men (OR: 0.91, 95% CI 0.87 to 0.94). In women, higher red and processed meat intake was associated with higher odds of infertility (adjusted OR per 1-SD increment: 1.04, 95% CI 1.02 to 1.07), and a nominal association was observed for the snack/fast food proxy (OR 1.03, 95% CI 1.01 to 1.05). Whole grains and fish showed evidence of non-linear associations: the lowest estimated odds of infertility were observed at approximately 5 servings/day for whole grains, and at approximately 0.4 serving/day for fish. In men, higher intakes of fruits, vegetables, whole grains, and fish were associated with lower odds of infertility (5-6% lower odds per 1-SD higher intake). Higher values of the snack/fast food proxy were associated with higher odds of infertility (7% higher odds per 1-SD higher intake). Associations were attenuated in women after adjustment for their partner’s modified DASH score, but not vice versa.

**Conclusion:** Greater adherence to a modified DASH dietary pattern was associated with lower odds of infertility in both women and men, with stronger and more consistent associations in men and partly sex-specific food-group patterns, although maternal diet was assessed during pregnancy and may not reflect preconception intake. These findings support considering overall diet quality and both partners’ diets when studying infertility and highlight the need for preconception studies and dietary interventions using clinically relevant couple-level fertility outcomes.

## 1. INTRODUCTION

Infertility is defined as failure to achieve pregnancy after at least 12 months of regular unprotected intercourse. It affects almost 1/5 of couples and is increasingly recognized as a condition with medical, psychosocial, and public-health relevance [1,2]. Infertility may result from male or female reproductive factors, a combination of both, or remain unexplained [3]. Modifiable cardiometabolic and behavioral factors, including body mass index, impaired glucose metabolism, and smoking, may contribute [4–7].

Diet is also a plausible determinant of fertility, potentially influencing ovulatory function, endocrine regulation, sperm quality, the follicular and seminal environments, and systemic inflammation [8–10]. In women, higher intakes of fruits, vegetables, whole grains, and seafood have generally been associated with better fertility-related outcomes, whereas trans and saturated fats and added sugars have been associated with poorer outcomes [8–11]. In men, evidence relates mainly to semen quality. A higher intake of fruits, vegetables, whole grains, nuts, low-fat dairy products, and seafood, and a lower intake of red and processed meat, sweets, and sugar-sweetened beverages has been associated with more favorable semen quality parameters [10,12,13]. Evidence on how diet may impact clinical infertility remains limited, particularly in men. Previous findings are largely derived from smaller studies restricted to couples undergoing fertility treatment, and these studies have generally assumed linear associations between dietary intakes and infertility [8–10]. Few studies have examined the association between overall diet quality and infertility using established healthy dietary patterns, such as the Dietary Approaches to Stop Hypertension (DASH) diet. This dietary pattern emphasizes fruits, vegetables, whole grains, nuts, and low-fat dairy products while limiting red and processed meat and sugar-sweetened beverages [14], and has been associated with favorable cardiometabolic profiles [15]. Thus, we aimed to investigate whether adherence to a healthy dietary pattern assessed using a modified DASH score, and intake of 9 key food groups (fruits, vegetables, nuts/legumes, whole grains, sweet beverages, total dairy, fish, red and processed meat, and snacks) were associated with infertility in women and men.

## 2. MATERIALS AND METHODS

### 2.1. Study population

This study used data from The Norwegian Mother, Father and Child Cohort Study (MoBa), a population-based pregnancy cohort conducted by the Norwegian Institute of Public Health [16,17]. Pregnant women and their partners were recruited throughout Norway in connection with routine ultrasound examinations (around 18^th^ gestational week) between 1999 and 2008. MoBa involves approximately 114,500 children, 95,000 mothers, and 75,000 partners, and includes questionnaire data, biological samples, and linkage to the Medical Birth Registry of Norway. The analytic samples herein comprised unique mothers and male partners from planned pregnancies with non-missing infertility information and available sex-specific dietary data: 59,218 women with maternal dietary data and 26,866 male partners with paternal dietary data (**Figure 1**). Participants could contribute more than one eligible pregnancy, with dietary exposures and infertility status assessed for each pregnancy separately. Our work is described according to the Strengthening the Reporting of Observational studies in Epidemiology guidelines for cohort studies.

**Figure 1.**
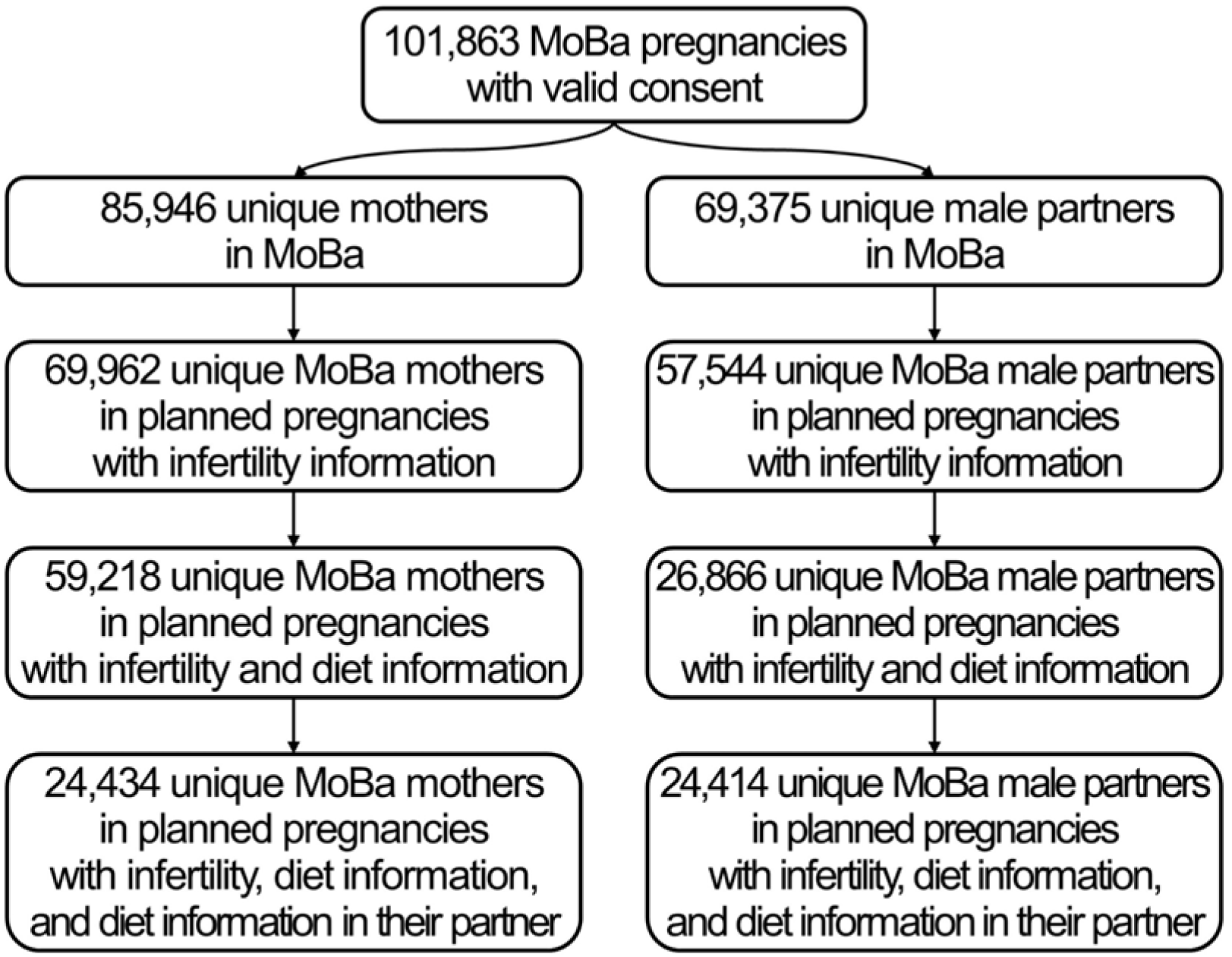
Flow chart of participant selection for the maternal and paternal analytic samples from the Norwegian Mother, Father and Child Cohort Study

### 2.2. Dietary assessment and modified DASH score

Dietary intake was assessed using MoBa questionnaires administered during pregnancy. The women self-reported dietary intake through the validated MoBa 255-item semi-quantitative food-frequency questionnaire, administered around gestational week 22 and covering dietary intake since the beginning of pregnancy [18,19]. Male partners completed a shorter self-reported questionnaire around gestational week 18 covering bread and bread toppings, beverages, dinner meals, fruits and vegetables, and eating patterns, without a specified dietary reference period. Because maternal diet was assessed during pregnancy, it may partly reflect pregnancy-related dietary changes [20]. Participants with calculated maternal energy intakes <4.5 or >20 MJ/day were excluded [21].

In women, fruits included fresh fruits, dried fruits, fruit used as bread spread, and a fraction of fruit juice, with the first 200 g/day contributing up to 100 g/day to fruit intake and any intake above 200 g/day assigned to sweet beverages [22]. Vegetables included raw and cooked vegetables, vegetables used as bread toppings, vegetables served with main dishes, and the estimated vegetable fraction of composite dishes such as casseroles, soups, wok dishes, pasta dishes, pizza, tacos/burritos, and vegetable-based main dishes. Nuts/legumes, treated as a single component according to the DASH score [23], included peanuts, almonds/hazelnuts/cashews and other nuts, nut spreads, vegetarian spreads, beans, lentils, and soya-based dishes. Whole grains included whole meal products, dark, fiber-rich bread/crispbread and whole-grain cereals or porridge. The DASH low-fat dairy component was calculated using intakes of low-fat milk/yoghurt and low-fat cheese (other dairy products were not included because the DASH score specifically considers low-fat dairy [23]). The DASH sweet beverage component was calculated by all cordial/soft drinks, caffeinated soft drinks, and fruit juice exceeding 200 g/day. Red and processed meat included pork, beef/veal, lamb, game meat, offal, minced meat, processed meat products (meat-based bread spreads, sausages, meatballs, hamburgers, and bacon), and the estimated meat fraction of composite dishes. Because sodium intake was unavailable, a snack/fast food proxy was used in its place for the modified DASH score. The proxy was calculated by summing the reported daily frequencies of food purchased from kiosks/snack bars, petrol stations, and fast food restaurants. Finally, fish, which is not a component of the DASH score but was additionally assessed, included fish used as bread spreads; lean and fatty fish consumed as meals; fish cakes, fish pudding, fish balls, and breaded fish; and the estimated fish fraction of fish-containing casseroles, soups, and pasta dishes.

In men, food groups were derived from the shorter MoBa father questionnaire. Fruits included reported fruit intake and fruit juice, applying the same approach as in women: reported glasses/day were converted to grams/day, with the first 200 g/day contributing proportionally up to 100 g/day to fruit intake and any excess assigned to sweet beverages [22]. Vegetables included raw vegetables/salads, cooked vegetables in stews, and cooked vegetables. As nuts and legumes were not directly assessed, the frequency of vegetarian meals was used as a proxy and converted to approximate intake using a 150 g portion. Whole grains included medium coarse-grain bread and coarse-grain bread. The low-fat dairy component included low-fat/skimmed milk and reduced-fat cheese, using 250-g and 15-g portions, respectively. Sweet beverages included excess fruit juice, sugar-sweetened cola/soft drinks, non-sugar-sweetened cola/soft drinks, other sugar-free fizzy drinks, and energy drinks. Red and processed meat included lean meat used as bread topping, servelat/salami, liver pâté, sausages/hamburgers, kebab, meals with minced meat, pure meat, and the estimated meat contribution from pizza. Because sodium intake was unavailable, a snack/fast food proxy was used in its place for the modified DASH score, calculated by converting reported frequencies of food purchased from kiosks/snack bars, petrol stations, and fast food restaurants to daily frequencies and summing them. Fish included fish used as bread spread, lean fish, fatty fish, and fish balls/fish cakes.

The main exposure was overall diet quality, operationalized using a modified DASH diet score adapted from Fung et al. [23]. The score comprised eight components defined above: five encouraged components (fruits, vegetables, nuts/legumes, whole grains, and low-fat dairy) and three components to be limited (sweet beverages, red and processed meat, and the snack/fast-food proxy replacing sodium). Each component was ranked into sex-specific quintiles and assigned 1-5 points, with higher intakes of encouraged components receiving higher scores and higher intakes of components to be limited receiving lower scores. Component scores were summed to obtain a modified DASH score ranging from 8 to 40, with higher scores indicating greater adherence to a DASH-like dietary pattern. As secondary exposures, the eight food group variables contributing to the modified DASH score were examined separately, together with fish as an additional food group (nine food groups in total).

### 2.3. Infertility

During recruitment to MoBa, women reported whether the current pregnancy was planned and provided information on how long they had tried to conceive; initial response categories captured short waiting times (<1 month and 1-2 months), whereas attempts lasting ≥3 months were followed by a request for the exact number of months [16]. Infertility was defined among planned pregnancies as having tried to conceive for ≥12 months or having conceived after use of assisted reproductive technologies (ART). Information on use of ART was collected from the Medical Birth Registry, given that individuals offered ART in the public Norwegian health care system are expected to have tried to conceive for ≥12 months before referral [24]. Individuals who reported a planned pregnancy and achieved conception within 12 months of trying were classified as the reference group. For male-specific analyses, the infertility status registered for each pregnancy was attributed to the male partner linked to that pregnancy.

### 2.4. Other variables

For each pregnancy, we obtained information from both parents on their age (continuous), years of education (continuous), body mass index (pre-pregnancy for women; continuous), having ever smoked (yes/no), and number of offspring (0, 1, 2, or ≥ 3).

### 2.5. Ethical approval

MoBa is conducted in accordance with the principles of the Declaration of Helsinki for research involving human participants. The cohort was established and the first waves of data collection were carried out under a license from the Norwegian Data Protection Agency and with approval from the Regional Committees for Medical and Health Research Ethics; MoBa is currently regulated under the Norwegian Health Registry Act. All participants gave written informed consent before enrollment. The present study was approved by the Regional Committee for Medical and Health Research Ethics of South/East Norway (reference 2017/1362).

### 2.6. Statistical analyses

Participant characteristics were summarized at the participant level, using information from the first eligible pregnancy for age at delivery, education, body mass index, smoking, parity, and dietary exposures. Infertility and use of assisted reproductive technologies were summarized according to whether participants reported these outcomes in any eligible pregnancy. Continuous variables were summarized using means and standard deviations for normally distributed variables, medians and first and third quartiles for non-normally distributed variables, and proportions for categorical variables. To assess potential selection bias, participants with and without diet information in MoBa were compared using Student’s t tests for normally distributed continuous variables, Mann-Whitney U tests for non-normally distributed continuous variables, and chi-squared tests for categorical variables.

The primary analyses evaluated the association between modified DASH score and infertility using logistic regression models fitted separately in women and men, with results expressed as odds ratios (ORs) and 95% confidence intervals (CI) per 1-SD higher score. Secondary analyses evaluated the nine individual food groups separately in women and men. Food-group intakes were log-transformed as log(1 + intake) before analysis. For linear analyses, log-transformed intakes were standardized, with ORs expressed per 1-SD higher intake. Dose-response associations were examined using cubic spline models (4 degrees of freedom) fitted to the log-transformed intakes. Non-linearity was assessed by comparing linear and cubic spline models using likelihood ratio tests. When evidence of non-linearity was detected, spline models were used to characterize the shape of the association and identify the intake associated with the lowest odds of infertility. For presentation and interpretation, spline estimates were back-transformed to the original intake scale and food-group intakes were expressed as servings/day using predefined serving sizes: 100 g for fruits, vegetables, fish, and red and processed meat; 30 g for nuts/legumes; 40 g for whole grains; 200 g for low-fat dairy; and 250 g for sweet beverages. For the snack/fast food proxy, intake was expressed as the reported frequency of purchasing food from kiosks/snack bars, petrol stations, and fast-food restaurants. Zero intake, or the lowest observed intake when zero was not observed, was used as the reference. Models were fitted unadjusted and adjusted for age at delivery, years of education, body mass index, ever smoking, and number of offspring. To account for dietary concordance within couples, analyses were repeated among participants with dietary information available for both partners and further adjusted for the partner’s modified DASH score. Unlike the participant-level descriptive analyses, regression analyses included each eligible pregnancy as a separate observation, with dietary exposures, infertility status, and covariates assessed for the corresponding pregnancy. Cluster-robust standard errors accounted for repeated pregnancies contributed by the same participant. For food group analyses, multiple testing was addressed using Horn’s parallel analysis to estimate the effective number of independent comparisons (*m)* [25], followed by application of a Bonferroni-corrected significance threshold of 0.05/*m*. Analyses were performed in R Software version 4.0.3. Code for data management and statistical analysis has been made available in https://github.com/alvarohernaez/diet_infertility_moba.

## 3. RESULTS

### 3.1. Study population

The final analytic samples included 59,218 women and 26,866 men (**Figure 1**). These participants could contribute more than one eligible pregnancy to the regression analyses. Approximately 15% of participants reported infertility in at least one eligible pregnancy. Participant characteristics at the first eligible pregnancy, including modified DASH scores and food-group intakes, are presented in **Table 1**, together with the proportions ever reporting infertility or use of assisted reproductive technologies. MoBa participants with dietary information were older, had higher educational attainment, were less likely to have smoked, had lower parity, and were more likely to experience infertility than participants without dietary information (**Supplemental Table 1**).

**Table 1.**
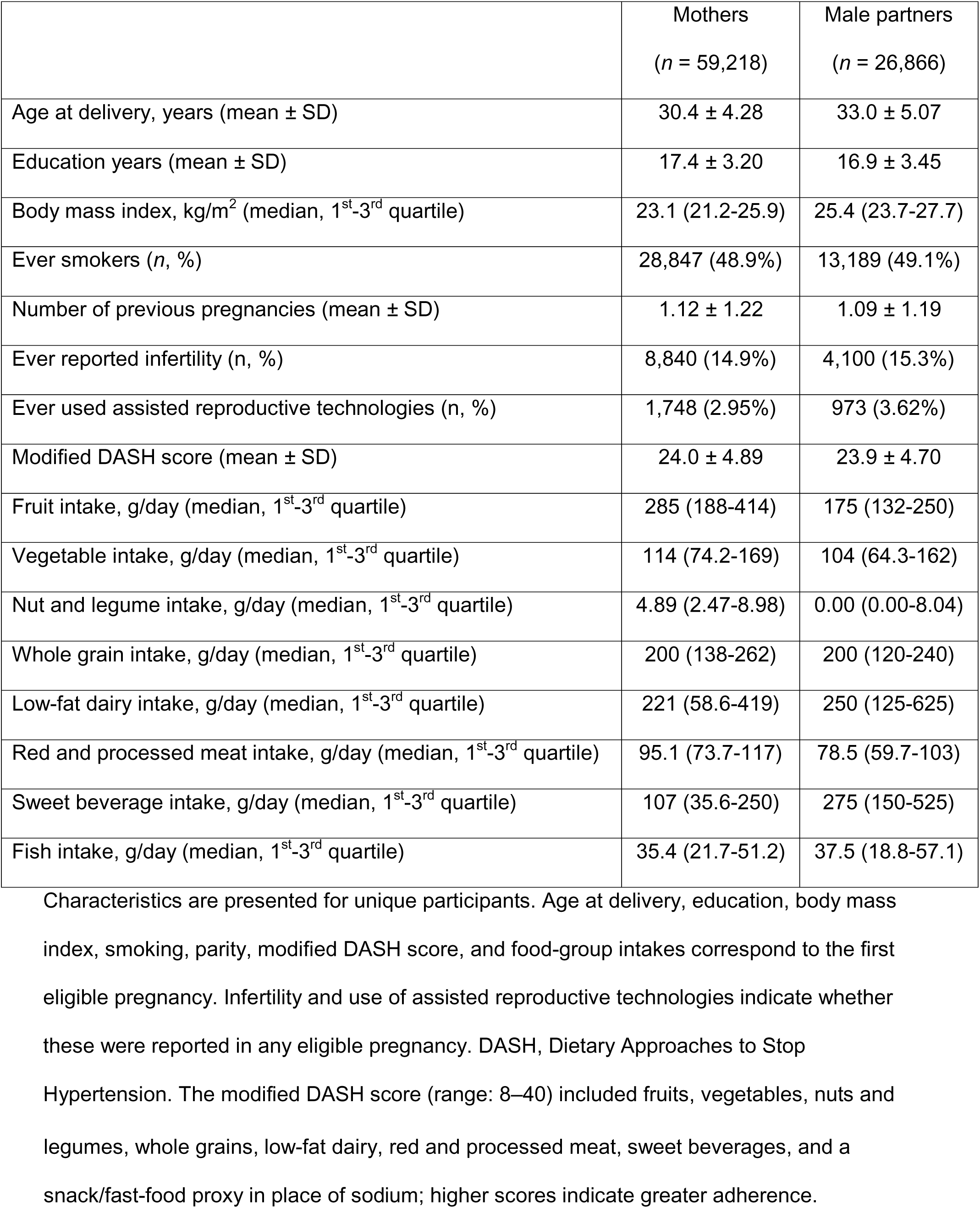
Characteristics of study participants.

Characteristics are presented for unique participants. Age at delivery, education, body mass index, smoking, parity, modified DASH score, and food-group intakes correspond to the first eligible pregnancy. Infertility and use of assisted reproductive technologies indicate whether these were reported in any eligible pregnancy. DASH, Dietary Approaches to Stop Hypertension. The modified DASH score (range: 8–40) included fruits, vegetables, nuts and legumes, whole grains, low-fat dairy, red and processed meat, sweet beverages, and a snack/fast-food proxy in place of sodium; higher scores indicate greater adherence.

### 3.2. Modified DASH score and infertility

Higher adherence to the modified DASH dietary pattern was associated with lower odds of infertility in both women and men (**Figure 2**), with adjusted ORs per 1-SD higher score of 0.97 (95% CI 0.95 to 0.995) in women and 0.91 (95% CI 0.87 to 0.94) in men. There was no evidence that these associations departed from linearity. The association in women was attenuated after adjustment for their partner’s modified DASH score, but this was not the case for the male participants.

**Figure 2.**
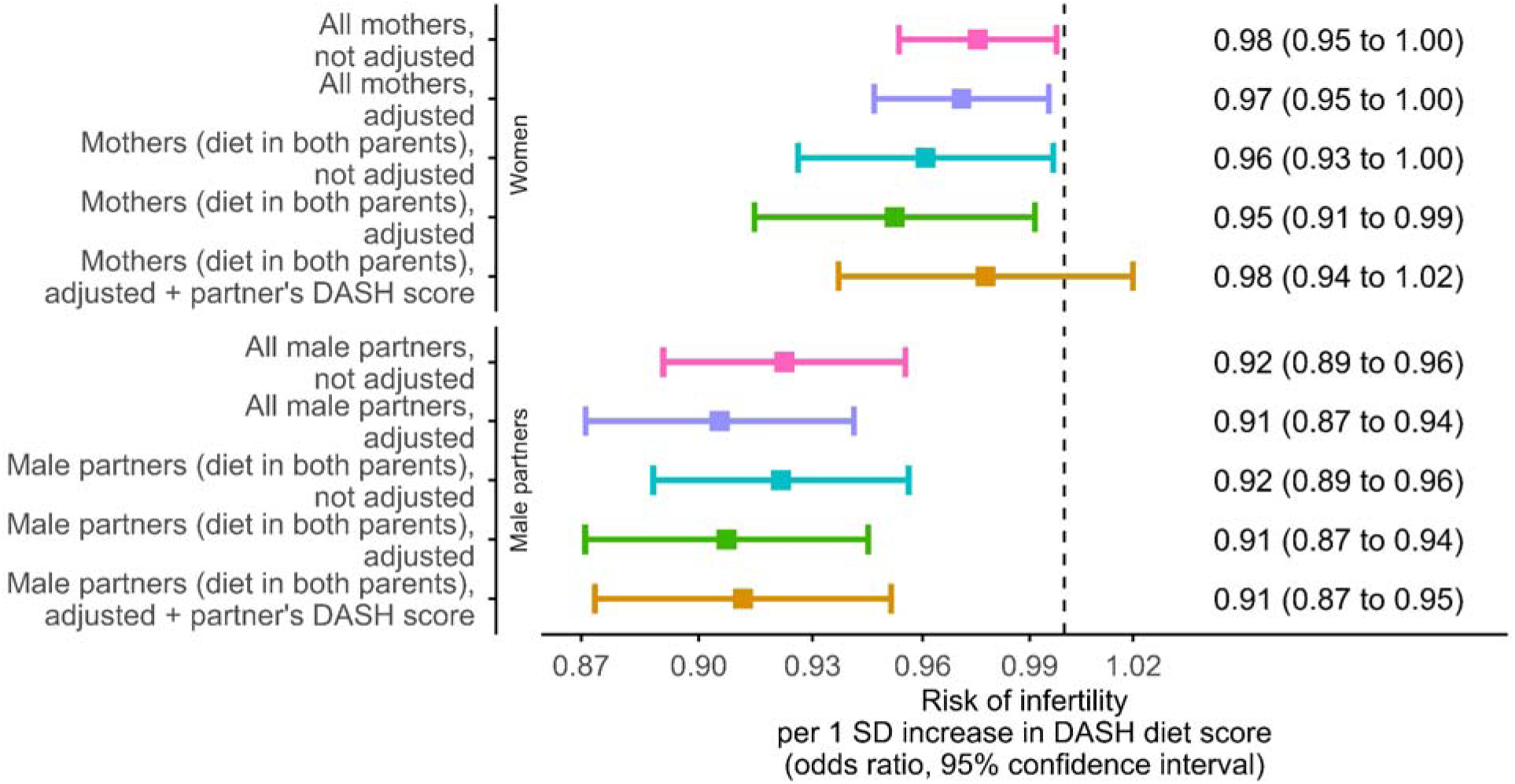
Association of a 1-SD higher score in the modified Dietary Approaches to Stop Hypertension diet with the odds of infertility (OR and 95% CI) in women (above) and male partners (below). DASH scores were standardized separately by sex. Plots show unadjusted associations among all women or male partners with dietary data (fuchsia); adjusted associations in the same samples (purple; adjusted for age at delivery, years of education, body mass index, ever smoking, and number of offspring); unadjusted associations restricted to women or male partners with dietary information available for both partners (emerald); adjusted associations in these restricted samples (green; same covariates); and associations further adjusted for the partner’s modified DASH score (orange).

### 3.3. Food groups and infertility in women

Horn’s parallel analysis suggested that three independent factors adequately captured the set of food group-related variables in women (multiple testing significance threshold: *p*-value < 0.0167). Intake of red and processed meat was linearly associated with greater infertility odds (per 1-SD higher intake: OR 1.04, 95% CI 1.02 to 1.07, *p*-value = 0.001, **Figure 3F**), whereas the proxy for snack/fast food intake was nominally associated with the outcome (per 1-SD higher intake: OR 1.03, 95% CI 1.01 to 1.05, *p*-value = 0.017) (**Figure 3H**). However, non-linear models better explained the associations of whole grains and fish with infertility (**Figures 3D** and **3I**): the lowest odds of infertility were observed at approximately 5 servings/day for whole grain intake (OR 0.85, 95% CI 0.83 to 0.87), and the first local minimum for fish intake occurred at approximately 0.4 servings/day (OR 0.88, 95% CI 0.86 to 0.90). Associations were attenuated or became more imprecise when adjusting for the partner’s modified DASH diet score (**Supplemental Figures 1A** and **2**).

**Figure 3.**
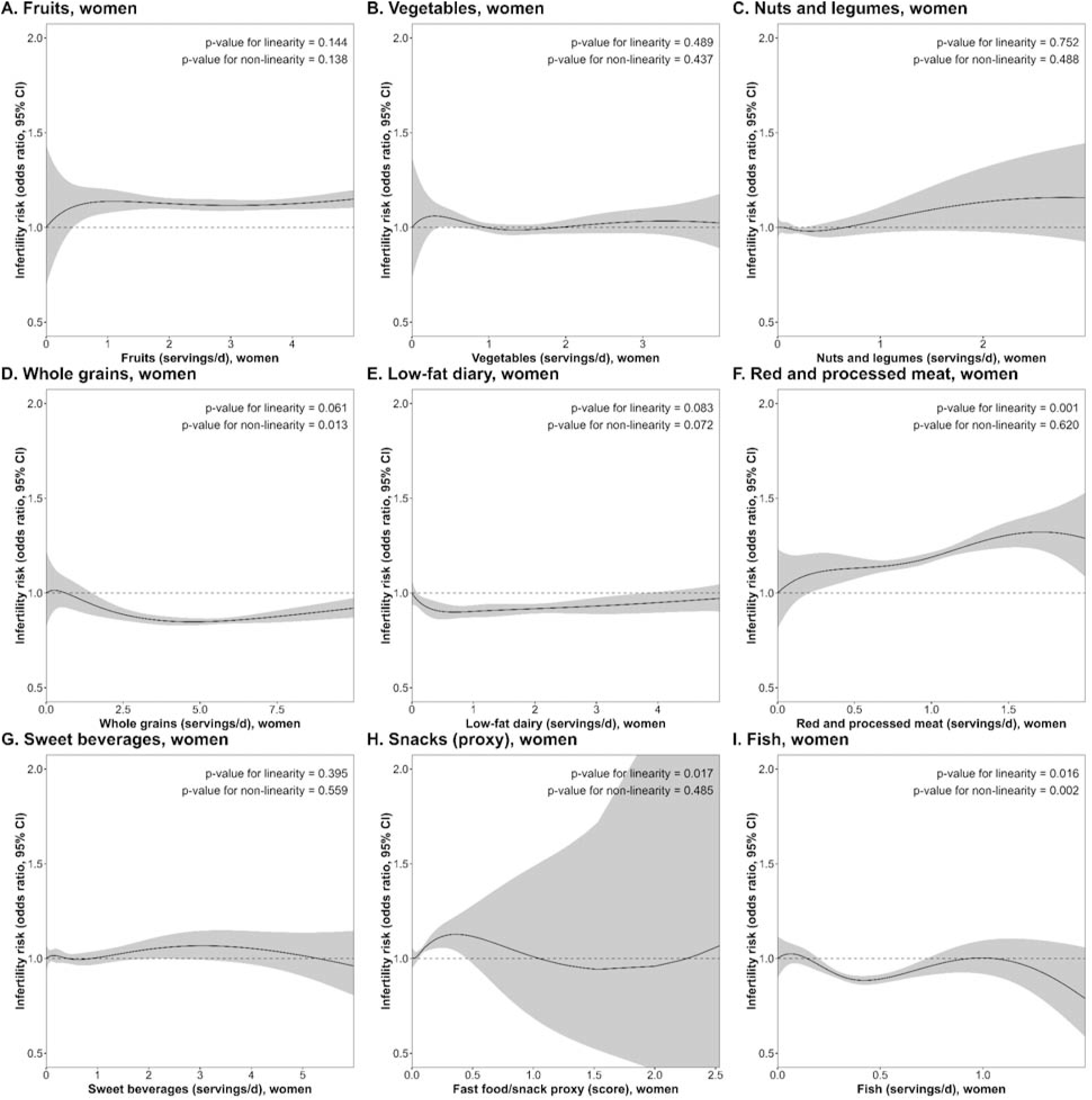
Associations of food group intakes (A: fruit; B: vegetables; C: nuts and legumes; D: whole grains; E: low-fat dairy; F: red and processed meat; G: sweet beverages; H: snack/fast food proxy; I: fish) with the odds of infertility (OR and 95% CI) in women. Associations were adjusted for age at delivery, years of education, body mass index, ever smoking, and number of offspring.

### 3.4. Food groups and infertility in men

Horn’s parallel analysis suggested that three independent factors adequately captured the set of food group-related variables in men (multiple testing significance threshold: *p*-value < 0.0167). Thus, in men (**Figure 4**), higher intakes of fruits (per 1-SD higher intake: OR 0.94, 95% CI 0.91 to 0.98, *p*-value = 0.001), vegetables (per 1-SD higher intake: OR 0.94, 95% CI 0.91 to 0.98, *p*-value = 0.002), the proxy for nut and legume intake (vegetarian dishes; per 1-SD higher intake: OR 0.94, 95% CI 0.91 to 0.98, *p*-value = 0.004), whole grains (per 1-SD higher intake: OR 0.94, 95% CI 0.91 to 0.97, *p*-value ≤ 0.001), and fish (per 1-SD higher intake: OR 0.95, 95% CI 0.92 to 0.99, *p*-value = 0.012) were associated with lower odds of infertility. A nominal association with low-fat dairy was also observed (per 1-SD higher intake: OR 0.96, 95% CI 0.93 to 1.00, *p*-value = 0.041). Conversely, higher values of the snack/fast food proxy were associated with higher odds of infertility (per 1-SD higher value: OR 1.07, 95% CI 1.03 to 1.11, *p*-value < 0.001) (**Figure 5**). There was no clear evidence that non-linear models improved model fit over linear models for any food group. Associations remained robust after adjustment for the partner’s modified DASH score (**Supplemental Figures 1B** and **3**).

**Figure 4.**
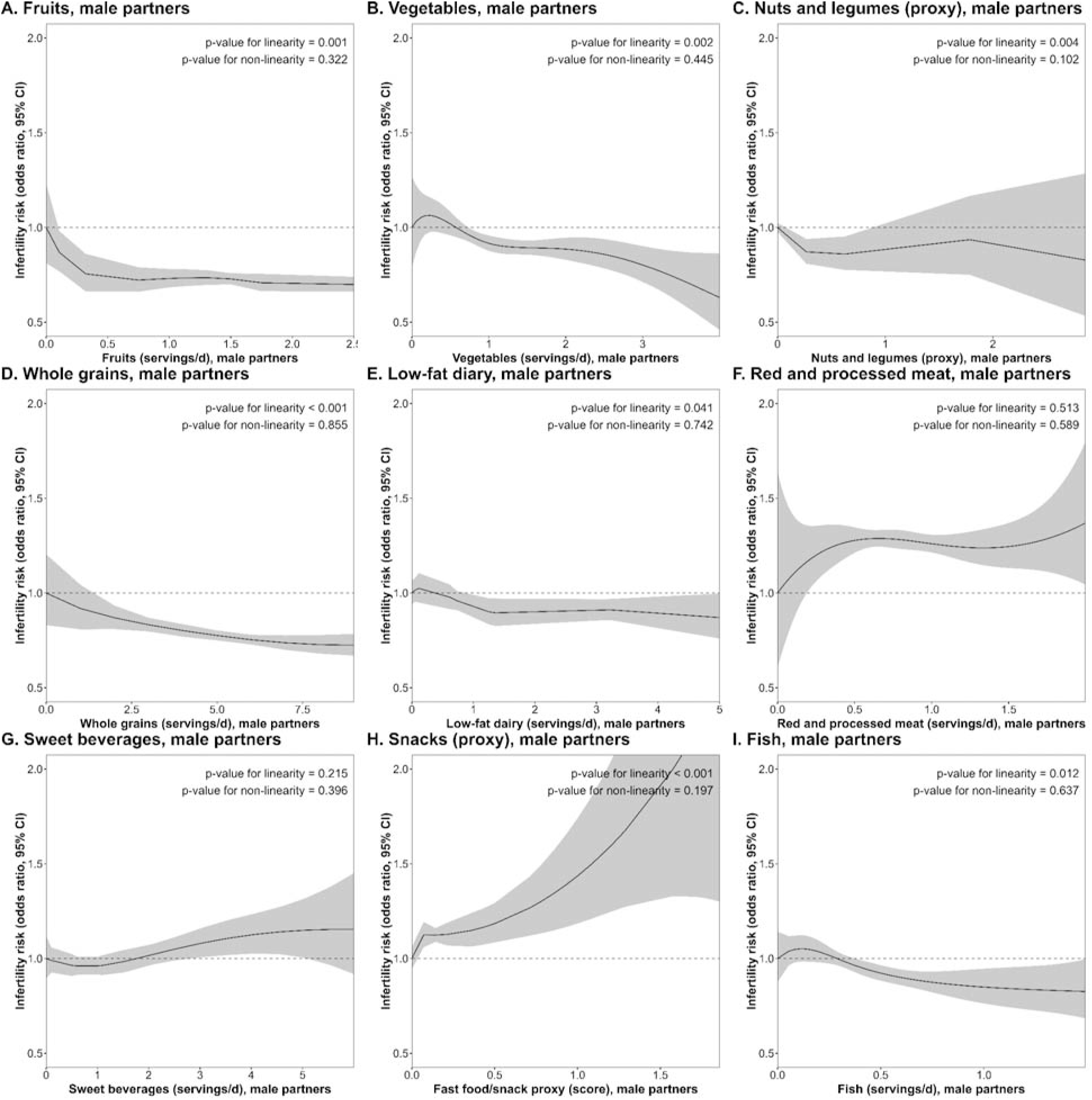
Associations of food group intakes (A: fruit; B: vegetables; C: nuts and legumes; D: whole grains; E: low-fat dairy; F: red and processed meat; G: sweet beverages; H: snack/fast food proxy; I: fish) with the odds of infertility (OR and 95% CI) in male partners. Associations are adjusted for age at delivery, years of education, body mass index, ever smoking, and number of offspring.

## 4. DISCUSSION

In this large population-based pregnancy cohort, higher adherence to a modified DASH diet was associated with lower odds of infertility in women and men. Secondary analyses suggested partly sex-specific food-group patterns. In women, higher red and processed meat intake was associated with higher odds of infertility, with a similar nominal association for the snack/fast food proxy, whereas lower odds were observed up to approximately 5 servings/day of whole grains and 0.4 servings/day of fish. In men, higher intakes of fruits, vegetables, a proxy for nuts and legumes (vegetarian dishes), whole grains, and fish were associated with lower odds of infertility, whereas higher snack/fast food proxy values were associated with higher odds of infertility. Associations were more numerous and generally stronger in men, while associations in women were attenuated after adjustment for their male partner’s dietary score; the converse was not observed for men.

### 4.1. Interpretation in women

Our findings in women are consistent with previous epidemiological evidence. A “fertility diet” pattern (prioritizing vegetable over animal protein, low glycemic load, full-fat dairy, iron, monounsaturated fats, and multivitamin use) was associated with lower risk of ovulatory disorder infertility [26], Mediterranean-style diets with lower odds of seeking medical help for difficulty conceiving [27], and higher-quality, less inflammatory diets with greater fecundability among pregnancy planners [11]. Higher red and processed meat intake has also been associated with poorer fertility outcomes [28,29]. The nominally higher odds of infertility associated with the snack/fast food proxy are consistent with evidence linking preconception fast-food intake to longer time to pregnancy and infertility [30]. Our results for whole grains are also broadly consistent with previous evidence: higher pretreatment whole-grain intake has been associated with a greater probability of live birth after ART [31], while higher dietary glycemic load has been associated with a higher infertility risk [32,33]. Similarly, higher fish and seafood intake in women and their partners has been associated with greater fecundity, shorter time to pregnancy [34] and a higher probability of live birth after ART [35]. Similar associations have been reported for women’s intake of omega-3-rich foods in relation to fecundability [36] and ART success [37]. However, fish-related contaminants, including mercury and polychlorinated biphenyls, have also demonstrated an adverse effect on reproductive outcomes [38–40]. The observed associations in women were generally modest and smaller than those in men, although their clinical relevance remains uncertain. Overall, these associations may reflect the potential reproductive benefits of higher intakes of fiber, antioxidants, omega-3 fatty acids, and other bioactive compounds, alongside reduced consumption of detrimental components of snacks and red and processed meats. These factors may support reproductive function through improved insulin sensitivity and ovulatory regulation, reduced inflammation and oxidative stress, and more favorable endocrine and follicular environments [6,8,33,41,42].

### 4.2. Interpretation in men

In male partners, our findings are broadly consistent with previous evidence, which is stronger for semen quality than for clinical fertility outcomes [10,13,43]. Although infertility is a couple-dependent outcome, our findings suggest that paternal diet may contribute to the likelihood of experiencing infertility. Higher adherence to healthy dietary patterns has been associated with better sperm quantity and quality [44–47]. However, evidence for clinical fertility outcomes is limited. Previous studies found no association between male overall diet quality and the likelihood of conceiving, while a study of healthy dietary patterns and ART outcomes (n=245 men) found no association with treatment success [48]. Our study extends this evidence to a clinically relevant infertility endpoint. Among individual dietary exposures, sugar-sweetened beverages have been associated with lower fecundability [49], and seafood with shorter time to pregnancy [34], whereas paternal dairy and omega-3 intake have not been associated with ART success [37,50]. Evidence from semen-related outcomes is more extensive, with associations reported for fruits and vegetables [51], low-fat dairy [52], fish [53], processed meat [53], soft drinks [54], and snack- and ultra-processed-food-rich diets [55]. These associations may reflect higher intakes of antioxidants, fiber, and omega-3 fatty acids and lower exposure to rapidly absorbed carbohydrates and detrimental components of processed meats, sweet beverages, and snacks. Such dietary characteristics may support male reproductive function by reducing oxidative stress, inflammation, and sperm DNA damage and preserving sperm membrane, mitochondrial, endocrine, and spermatogenic function [13,56–58].

### 4.3. Limitations

Our study has several limitations. First, infertility is a couple-dependent outcome and was defined from maternal time-to-pregnancy information and registry-based ART data; therefore, we could not distinguish female-factor, male-factor, combined, or unexplained infertility. Second, MoBa included only couples who ultimately conceived, mostly resulting in live births, limiting inference to couples with sterility. Third, our analyses are observational and residual confounding remains possible despite adjustment for age, education, body mass index, smoking, and parity, because diet quality is embedded within broader socioeconomic, behavioral, and health-related patterns. Therefore, causal associations between diet and fertility problems cannot be inferred. Fourth, we selected the DASH score as the most comprehensive dietary quality index that could be calculated consistently in women and men, given the limited detail of paternal dietary data and the minimal modifications required to its original scoring criteria. However, replacing sodium intake with a snack/fast food proxy may have introduced misclassification, and alternative dietary indices could yield different estimates of the associations with infertility. Fifth, there are several sources of potential exposure misclassification specific to women in our data. Questions on maternal diet reflected dietary patterns during pregnancy rather than during the preconception period (the father’s questionnaire did not specify a time period and may therefore better reflect usual diet), and women may have changed their diets after conception, either toward healthier choices or because of nausea, food aversions, or other pregnancy-related symptoms. Individuals experiencing infertility may also be more likely to adopt a healthier diet before conception, potentially leading to underestimation of the associations. The paternal dietary questionnaire was not validated and was less detailed than the maternal food frequency questionnaire, and one component of the modified DASH diet score was operationalized differently in men (vegetarian dishes as a proxy for nut and legume intake) due to the lack of direct information. These differences in dietary assessment may limit the direct comparability of association estimates between women and men. Repeated dietary assessments before and during pregnancy are therefore warranted to determine whether the observed sex-specific differences reflect systematic information bias. Sixth, the associations were modest (3% and 9% lower odds of infertility per 1-SD higher modified DASH score in women and men, respectively), suggesting that the contribution of diet to infertility risk may be relatively small compared with other established lifestyle risk factors, such as smoking and excess body weight. Finally, MoBa participants may differ from the general Norwegian population and from populations with different dietary patterns, fertility-care access, or sociodemographic characteristics, limiting the generalizability of our findings to Scandinavian individuals of child-bearing age.

## 5. CONCLUSION

In MoBa, greater adherence to a modified DASH dietary pattern was associated with lower odds of infertility in both women and men. In women, the food-group findings were driven mainly by higher red and processed meat intake, potentially by snack intake, and by non-linear patterns for whole grains and fish. In men, a favorable dietary profile (higher intakes of fruits, vegetables, vegetarian dishes, whole grains, and fish, and lower intakes of snacks) was associated with lower odds of infertility, and associations showed larger effect sizes and were more robust than in women. Our findings extend previous literature in several ways because, to our knowledge, this is the first study to use a very large sample of women and male partners, apply a clinically meaningful definition of infertility, and examine diet comprehensively, including both overall diet quality and individual food groups, with systematic assessment of non-linearity. From a clinical nutrition perspective, the results support studying whole-diet approaches rather than isolated nutrients, and they reinforce the need to include male partners when evaluating modifiable predictors of clinical infertility. Future studies should replicate our findings in other preconception cohorts with detailed clinical infertility phenotyping and characterize substitution patterns between food groups. Observational findings should also be triangulated with quasi-experimental and causal inference approaches. Together, these efforts could inform the design of randomized dietary intervention trials assessing clinically relevant couple-level fertility outcomes, such as infertility or time to pregnancy.

## Acknowledgements

The authors are grateful to all families participating in MoBa and to the Norwegian Institute of Public Health for generating and maintaining MoBa data. This work was performed on the Services for sensitive data (TSD) facilities, owned by the University of Oslo, operated and developed by the TSD service group at the University of Oslo, IT-Department. This study does not necessarily reflect the position or policy of the Norwegian Research Council. CIBER de Enfermedades Cardiovasculares (CIBERCV) and CIBER de Fisiopatología de la Obesidad y Nutrición (CIBEROBN) are initiatives of the Instituto de Salud Carlos III (Madrid, Spain) and are financed by the European Regional Development Fund. S. Castro-Barquero has received support from Instituto de Salud Carlos III, through the competitive Sara Borrell fellowship (code CD24/00244) co-funded by the European Union.

## Author contributions

Conceptualization: Á.H. Methodology: Á.H. Formal analysis: Á.H. Investigation: M.C.M. Data curation: Á.H. Writing-original draft: Á.H. Writing-review and editing: I.H.C., T.G., M.M.P., S.C.-B., M.H.H., and M.C.M. Funding acquisition: M.C.M. Supervision: M.C.M. All authors read and approved the final manuscript.

## Data availability

The consent provided by MoBa participants does not allow individual-level data to be deposited in public repositories or shared through journals. Researchers wishing to access the data for replication purposes should contact. Access requires approval from the Regional Committee for Medical and Health Research Ethics in Norway and a data-access agreement with MoBa.

