## Supplemental Materials for "Adherence to a modified DASH diet, intake of key food groups, and infertility in women and men: the Norwegian Mother, Father and Child Cohort Study"

**Supplemental Table 1.** Comparison of MoBa participants included and non-included in our analyses.

|  | Women | | | Male partners | | |
| --- | --- | --- | --- | --- | --- | --- |
|  | Included | Non-included | *p*-value | Included | Non-included | *p*-value |
| Age at delivery, years (mean ± SD) | 30.8 ± 4.3 | 29.7 ± 5.3 | <0.001 | 33.1 ± 5.1 | 32.8 ± 5.5 | <0.001 |
| Education years (mean ± SD) | 17.4 ± 3.2 | 16.2 ± 3.6 | <0.001 | 16.9 ± 3.4 | 15.9 ± 3.6 | <0.001 |
| Body mass index, kg/m^2^ (median, 1^st^-3^rd^ quartile) | 23.2  (21.3-26.1) | 23.1  (21.0-26.1) | <0.001 | 25.5  (23.7-27.8) | 25.5  (23.7-27.8) | 0.111 |
| Ever smokers (*n*, %) | 28,192  (49.4%) | 15,778  (57.4%) | <0.001 | 13,438  (50.3%) | 22,220  (53.2%) | <0.001 |
| Number of previous pregnancies (mean ± SD) | 1.3 ± 1.2 | 1.4 ± 1.4 | <0.001 | 1.1 ± 1.2 | 1.4 ± 1.3 | <0.001 |
| Infertility (*n*, %) | 8,493  (14.8%) | 1,988  (7.2%) | <0.001 | 4,084  (15.3%) | 4,433  (10.6%) | <0.001 |
| Use of assisted reproductive technologies (*n*, %) | 1,681  (2.9%) | 315  (1.1%) | <0.001 | 968  (3.6%) | 727  (1.7%) | <0.001 |

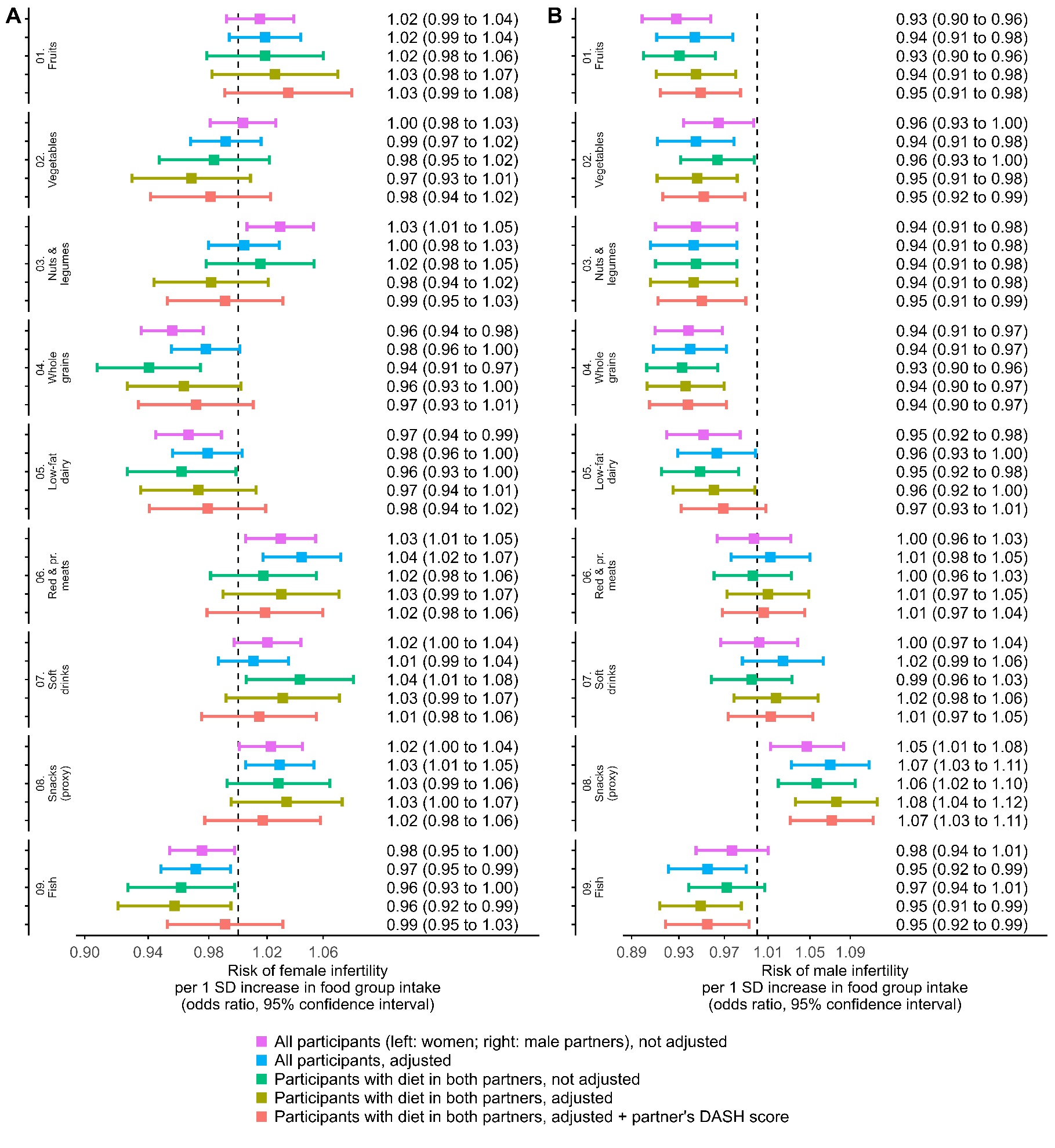
**Supplemental Figure 1.** Association of a 1-SD higher intake of food groups with the odds of infertility (OR and 95% CI) in women (A) and male partners (B). Food group intake values were standardized separately by sex. Plots show unadjusted associations among all women or male partners with dietary data (purple); adjusted associations in the same samples (blue; adjusted for age at delivery, years of education, body mass index, ever smoking, and number of offspring); unadjusted associations restricted to women or male partners with dietary information available for both partners (green); adjusted associations in these restricted samples (olive; same covariates); and associations further adjusted for the partner’s modified DASH score (orange).

**Supplemental** **Figure 2.** Associations of food group intakes (A: fruit; B: vegetables; C: nuts and legumes; D: whole grains; E: low-fat dairy; F: red and processed meat; G: sweet beverages; H: snack/fast food proxy; I: fish) with the odds of infertility (OR and 95% CI) in women with dietary information available for both partners. Associations were adjusted for age at delivery, years of education, body mass index, ever smoking, number of offspring, and the partner’s modified DASH score.

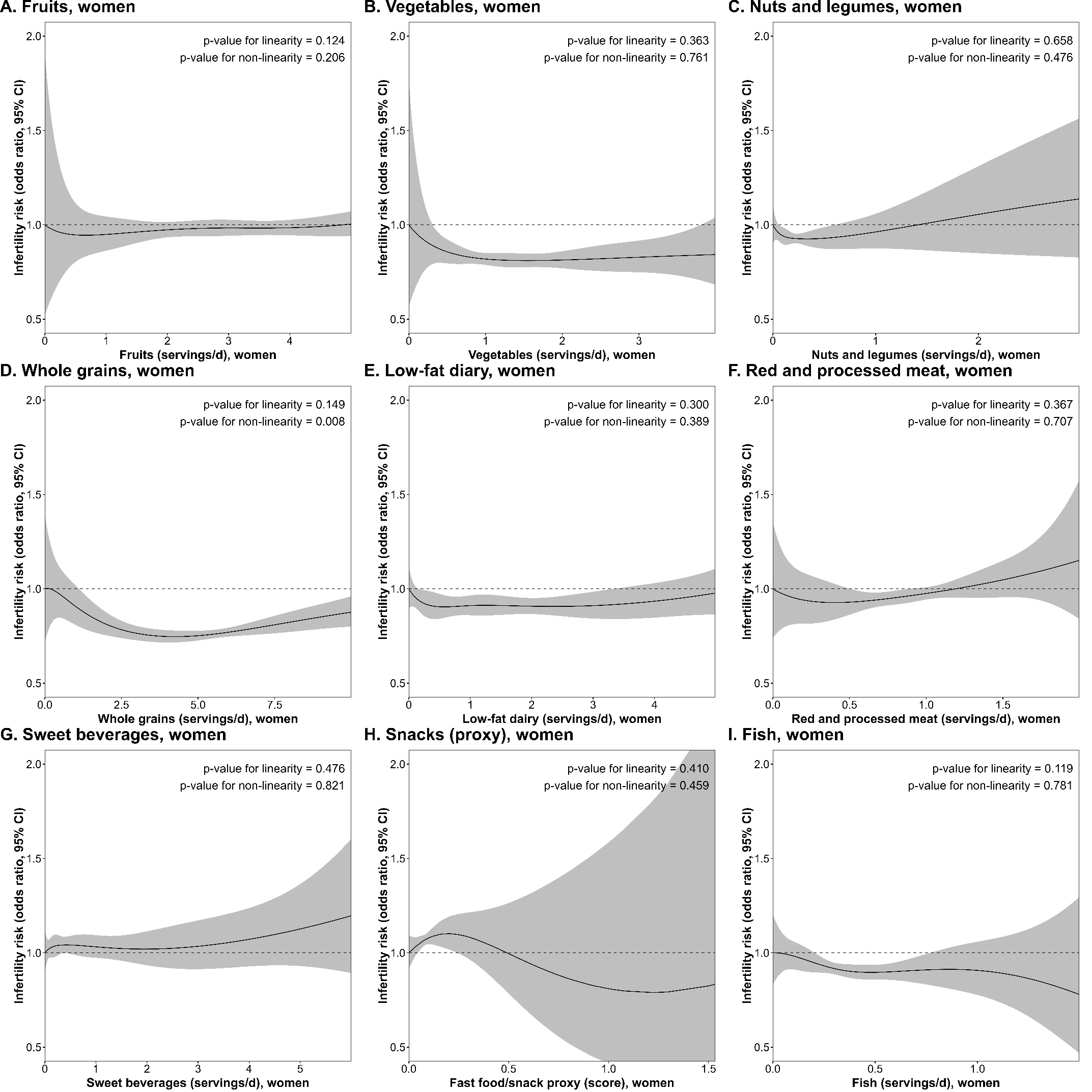

**Supplemental** **Figure 3.** Associations of food group intakes (A: fruit; B: vegetables; C: nuts and legumes; D: whole grains; E: low-fat dairy; F: red and processed meat; G: sweet beverages; H: snack/fast food proxy; I: fish) with the odds of infertility (OR and 95% CI) in male partners with dietary information available for both partners. Associations were adjusted for age at delivery, years of education, body mass index, ever smoking, number of offspring, and the partner’s modified DASH score.

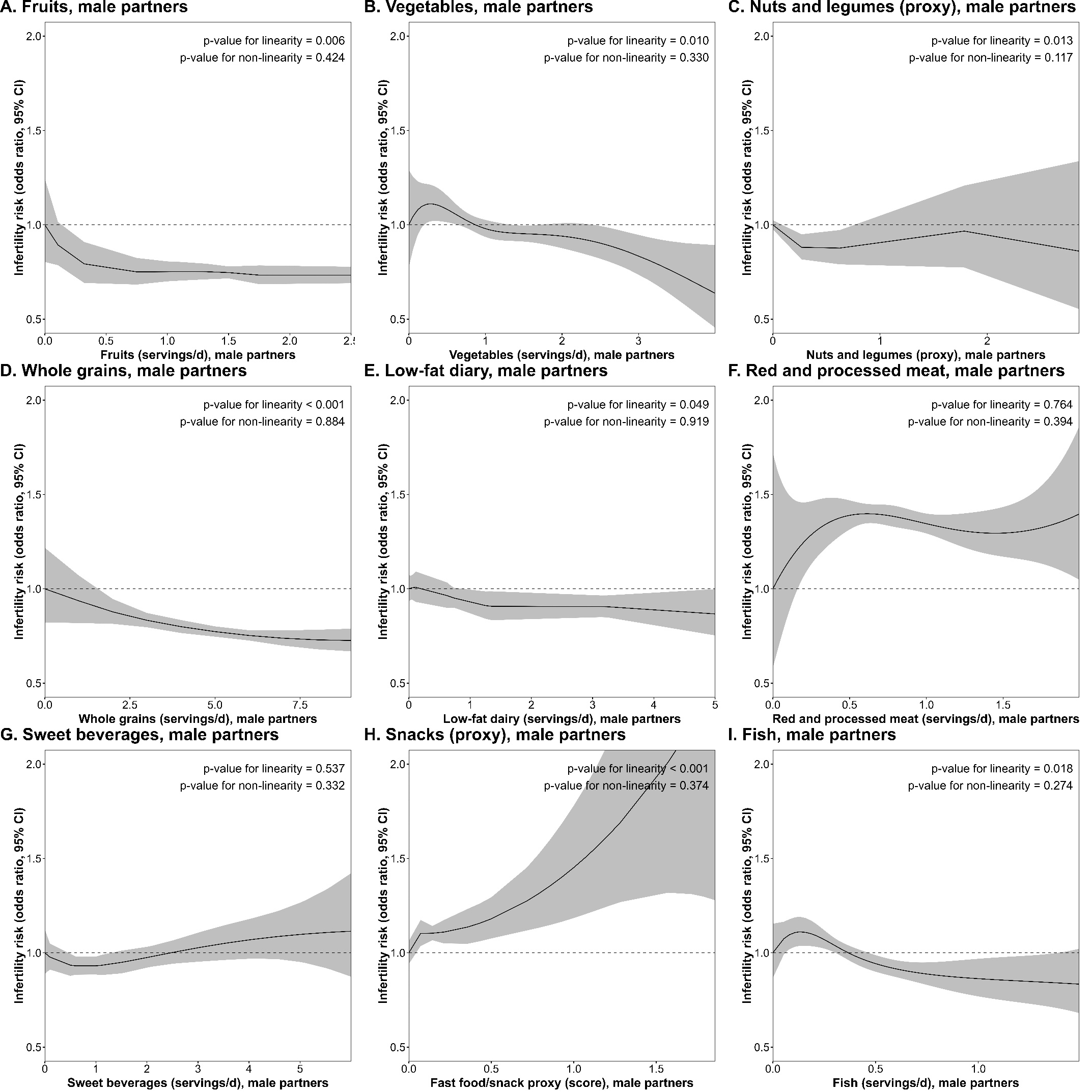
